# Preterm delivery and placental pathology with clinical and pathogenic implications

**DOI:** 10.64898/2026.04.09.26350526

**Authors:** Peilin Zhang

**Affiliations:** Pathology, Sharp HealthCare Laboratories, Pacific Rim Pathology

**Keywords:** Preterm delivery, placental pathology, maternal vascular malperfusion, preeclampsia, intrauterine fetal death, intrauterine infection, intra-amniotic infection, chorioamnionitis

## Abstract

**Background:** Preterm birth is one of the most significant etiologies for neonatal morbidity and mortality. Preterm delivery is classified as iatrogenic preterm delivery and spontaneous preterm delivery. The role of placental pathology is studied.

**Materials and methods:** We have previously collected placental pathology data with maternal pregnancy and neonatal birth data, and we investigated the role of placental pathology in preterm delivery. Preterm delivery was categorized as late preterm (34-36 weeks), moderate preterm (32 to 33 weeks), and extreme preterm (less than 32 weeks). Neonatal, maternal, placental gross and histologic features, and laboratory parameters were compared across groups using chi-square tests for categorical variables and Kruskal-Wallis tests for continuous variables using various programs in R-package.

**Results:** Totally 3723 singleton placentas including 3307 term (88.8%) and 416 preterm placentas (11.2%) were examined with maternal pregnancy data and neonatal birth data. There were 614 placentas from patients with preeclampsia/pregnancy induced hypertension (PRE/PIH) (16.5%). Preterm delivery showed significantly lower fetal birth weight, placental weight, and fetal-placental ratio (all p<0.01). Maternal Black race was more prevalent in preterm groups (up to 50.8% in extreme preterm vs. 33.2% in term, p<0.01). Preterm delivery was statistically associated with PRE/PIH and maternal vascular malperfusion (MVM), maternal and fetal inflammatory response (MIR and FIR), and increased pre-delivery white blood count (WBC). Extreme preterm deliveries were markedly associated with intrauterine fetal death (27.5%, p<0.01) and MIR/FIR (56.7%, p<0.01). After excluding PRE/PIH patients, preterm delivery was statistically associated with MIR/FIR and increased WBC.

**Conclusions:** Distinct clinicopathologic profiles exist across preterm subcategories, with MVM predominating in late/moderate preterm and severe pathologic features (including fetal demise and acute inflammation) in extreme preterm. These findings highlight heterogeneous etiologies of preterm delivery.

## Introduction

Preterm delivery is defined as birth before 37 weeks of gestation, and it affects approximately 10–12% of pregnancies worldwide ^1^. Preterm delivery is a major contributor to neonatal morbidity, mortality, and long-term neurodevelopmental disability. The etiology of preterm delivery is multifactorial, involving maternal, fetal, and placental factors. Preterm delivery has been classified clinically as late preterm (34–36 weeks), moderate preterm (32–33 weeks), and extreme preterm (<32 weeks), and these subcategories often reflect different underlying pathogenic mechanisms.

Placental examination provides critical insights into these pathogenic mechanisms^2^. Maternal vascular malperfusion (MVM) is frequently associated with hypertensive disorders of pregnancy such as preeclampsia/pregnancy induced hypertension (PRE/PIH), while fetal vascular malperfusion (FVM), acute inflammatory lesions including maternal and fetal inflammatory response (MIR/FIR), and chronic villitis may point to other pathways such as cord compromise, ascending infection (intra-amniotic infection), or immune-mediated processes ^3,4^.

This retrospective study describes neonatal, maternal, gross and histologic placental pathology, and laboratory features across term and preterm delivery categories in a large cohort of 3,723 cases with systematic placental evaluation. The aim was to identify clinicopathologic associations that may inform etiology and risk stratification.

## Materials and methods

The placental data collection was approved by the Institutional Review Board of New York Presbyterian – Brooklyn Methodist Hospital [1592673-1] (approval date 4-13-2020). The study included all term singleton placentas submitted chronologically for pathology examination from March 2020 and November 2021 except for twin or multiple births. Placental examination at this institution is criteria-based and performed according to the standard procedure. Placental pathology data, neonatal birth data, and maternal pregnancy data were retrieved from the medical records system (Cerner Corporation). The placental weight was measured after the fetal membrane and umbilical cord were trimmed. Only singleton pregnancies were included. Some of these data were previously published, but the issue of preterm delivery was not analyzed ^5-8^. The cases were categorized by gestational age at delivery: Term: ≥37 weeks (n=3,307), Late preterm: 34–36 weeks (n=258), Moderate preterm: 32–34 weeks (n=38), Extreme preterm: <32 weeks (n=120). Statistical analysis was performed by using various programs in R-package. Categorical variables were presented as counts (percentages) and compared using chi-square or Fisher’s exact tests. Continuous variables were presented as median [interquartile range] and compared using Kruskal-Wallis tests. A two-sided p-value <0.05 was considered statistically significant.

## Results

### 1, Preterm delivery and clinical pathologic features in entire cohort

A total of 3723 placentas with maternal pregnancy history, fetal birth data, and placental pathology were reviewed (Table 1). The entire case cohort was divided as term (n=3307), late preterm (n=258), moderate preterm (n=38), and extreme preterm (n=120) as defined previously. As expected, preterm deliveries were statistically significantly associated with progressively lower neonatal birth weight, head circumference, placental weight, and FPR (p<0.01 for all, Table 1). Ethnic black was significantly more common with preterm deliveries compared with other ethnic groups (46.1-50.8% vs 33.2%, p<0.01). Preterm deliveries were statistically associated with preeclampsia/pregnancy induced hypertension, diabetes, placental abruption and IUFD (all p<0.01). In placental histopathology, maternal vascular malperfusion (MVM) was significantly more common in preterm deliveries (Table 2, 14.0-18.4% vs 6.4%, p<0.01). Maternal inflammatory response (acute chorioamnionitis) and fetal inflammatory response (FIR) were significantly higher in prevalence in extreme preterm deliveries (both p<0.01). Chronic villitis and meconium-stained fetal membranes were significantly less common in preterm deliveries (both p<0.01). The white blood count (WBC) at the time of delivery was significantly higher in preterm deliveries with significantly elevated systolic blood pressure and body temperature (all p<0.01).

**Table 1:**
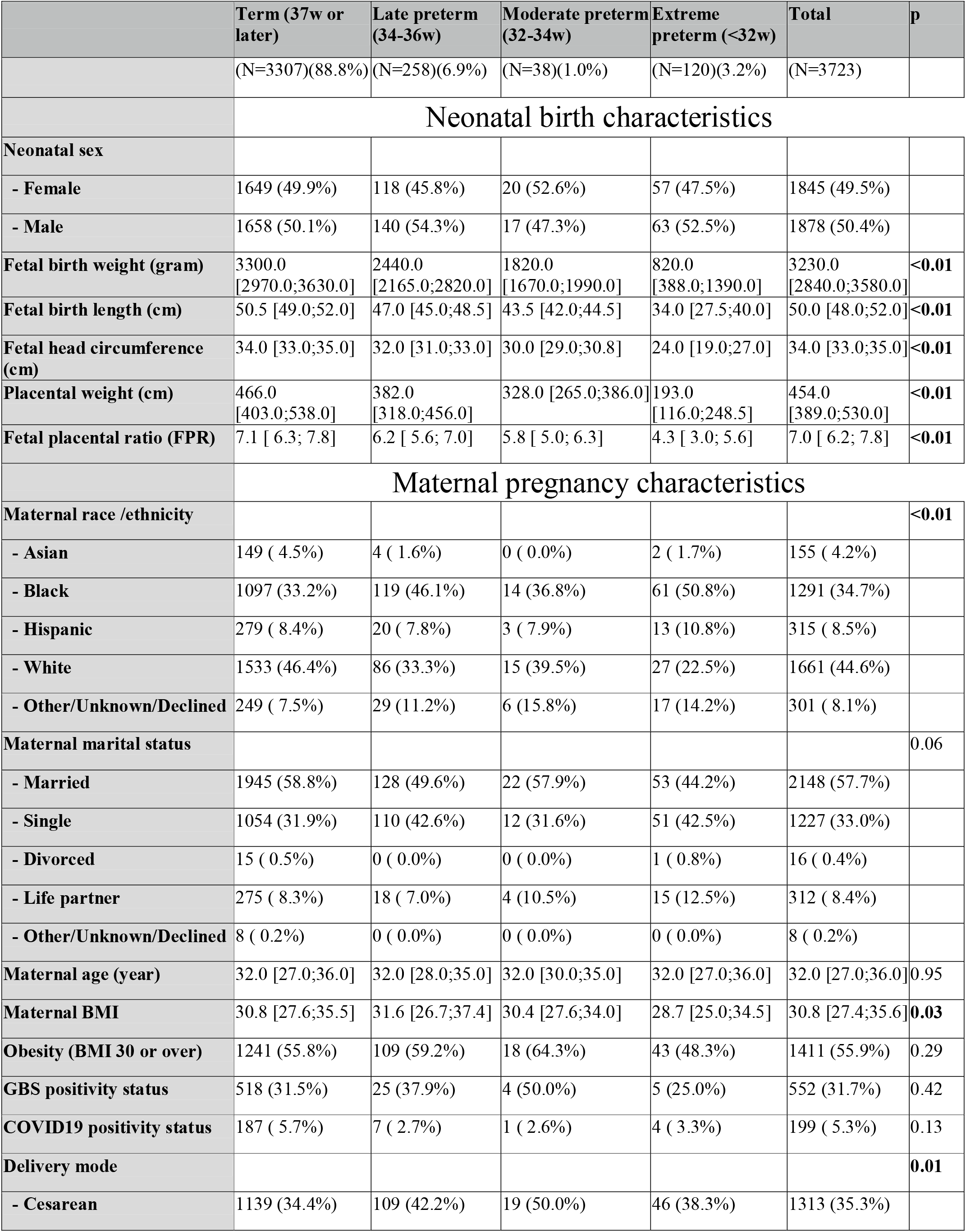
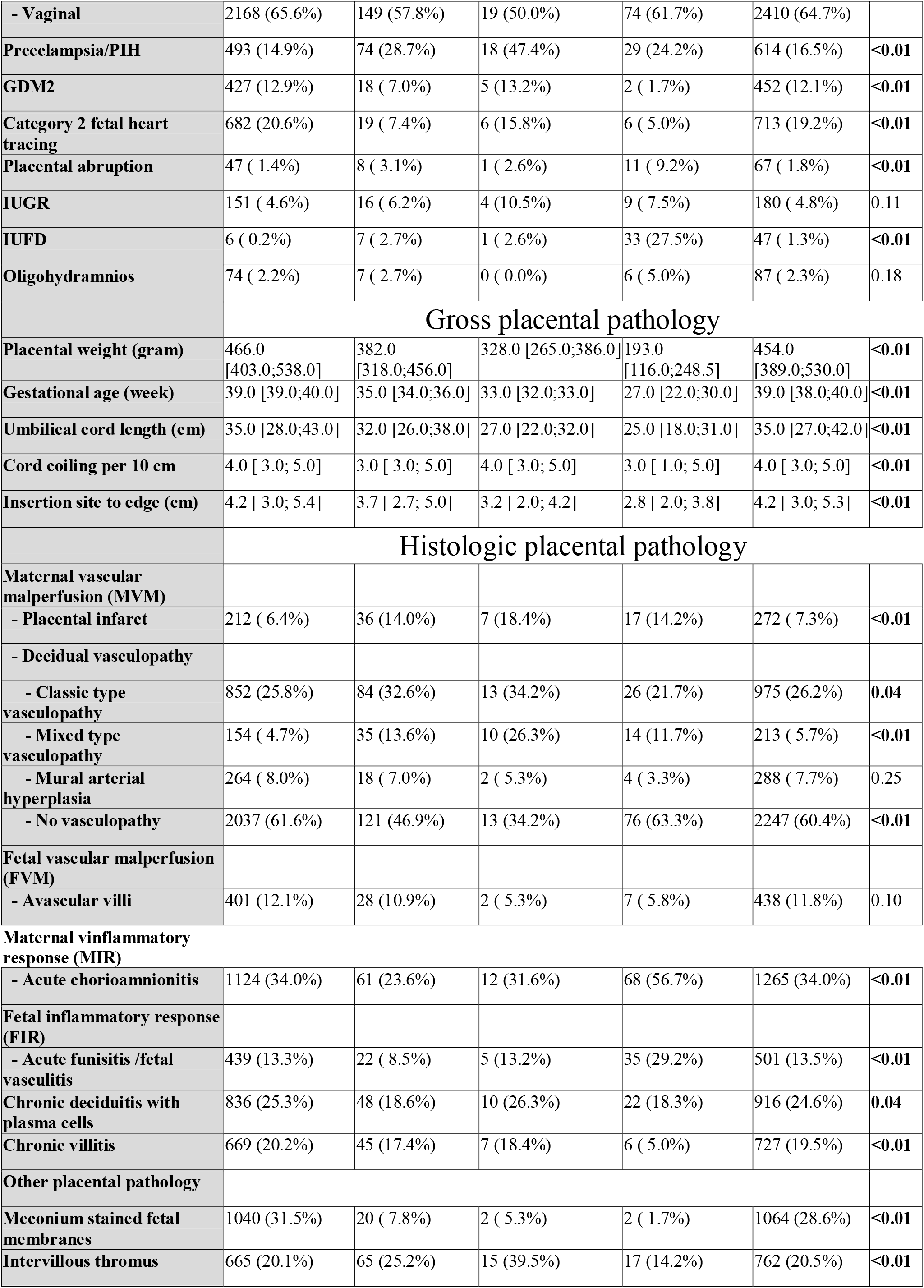
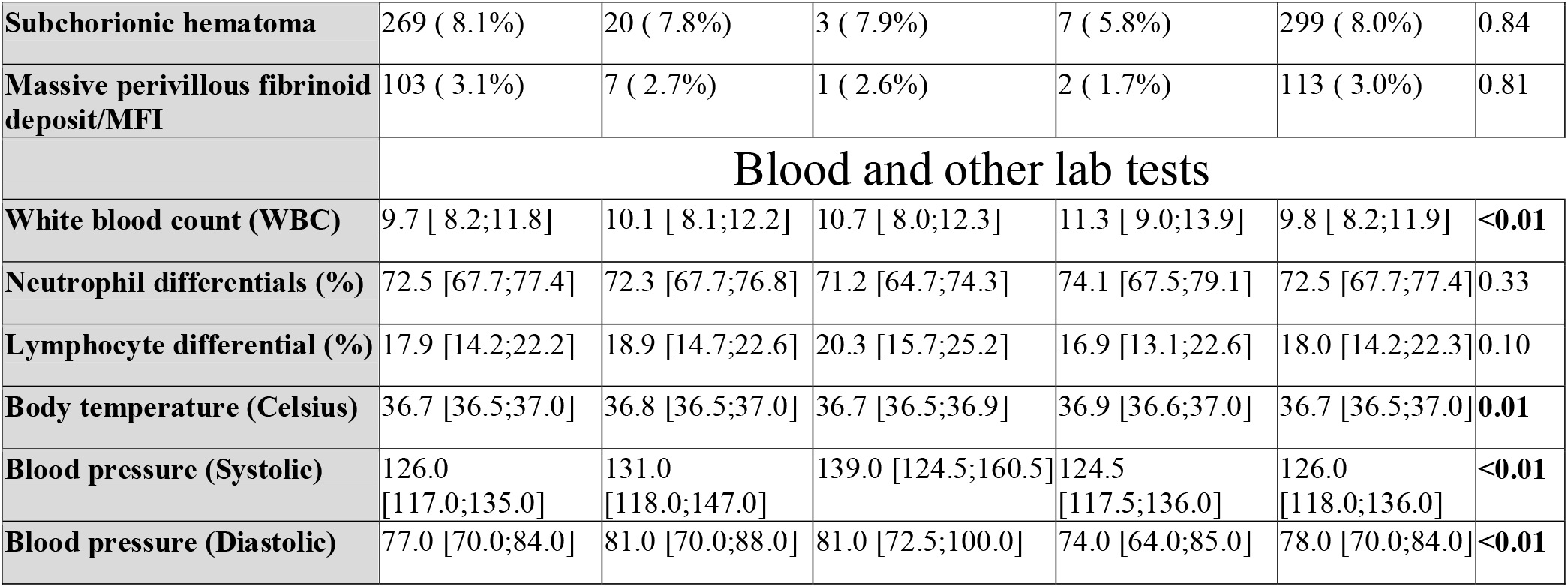
Preterm delivery and clinicopathologic features.

**Table 2:**
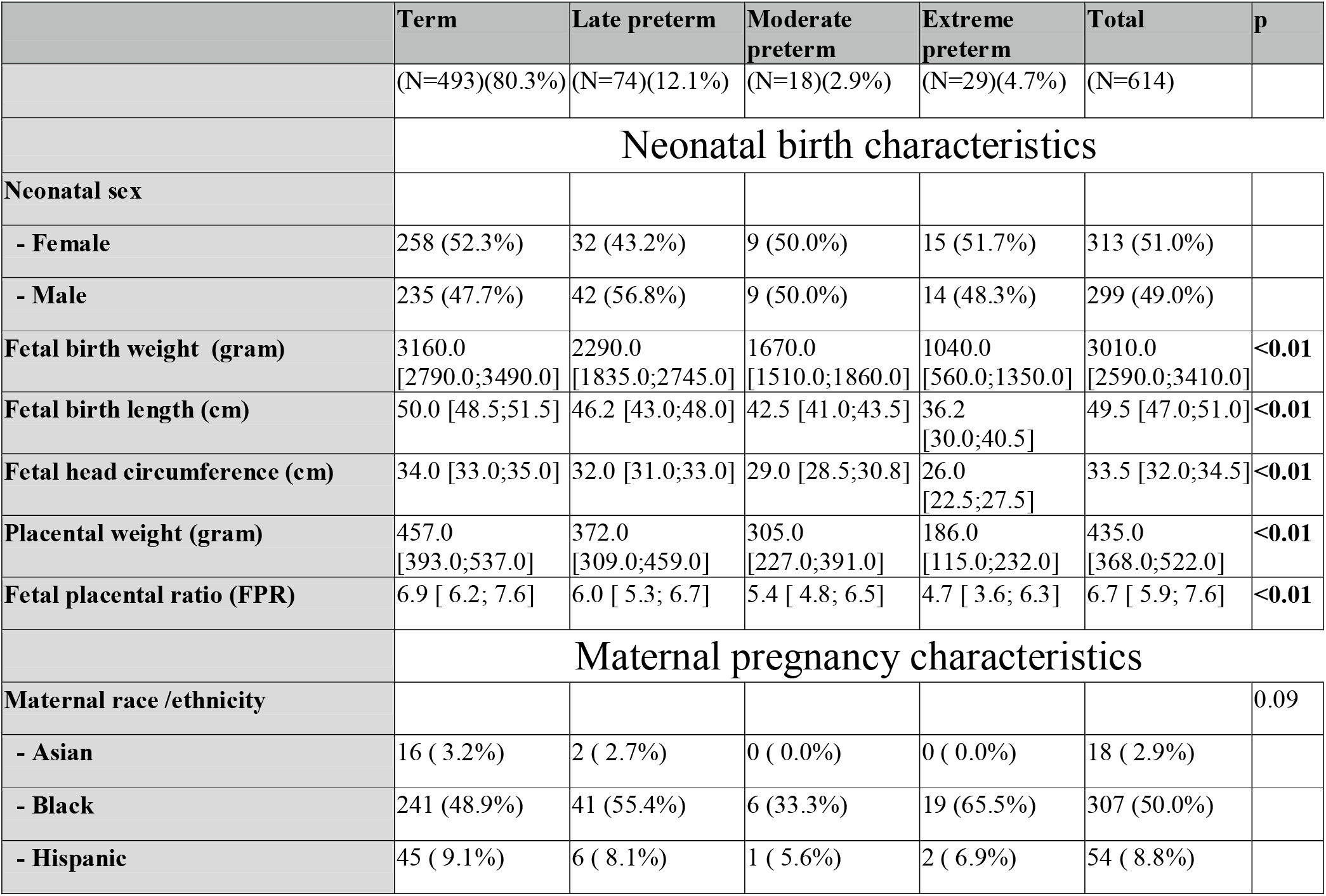
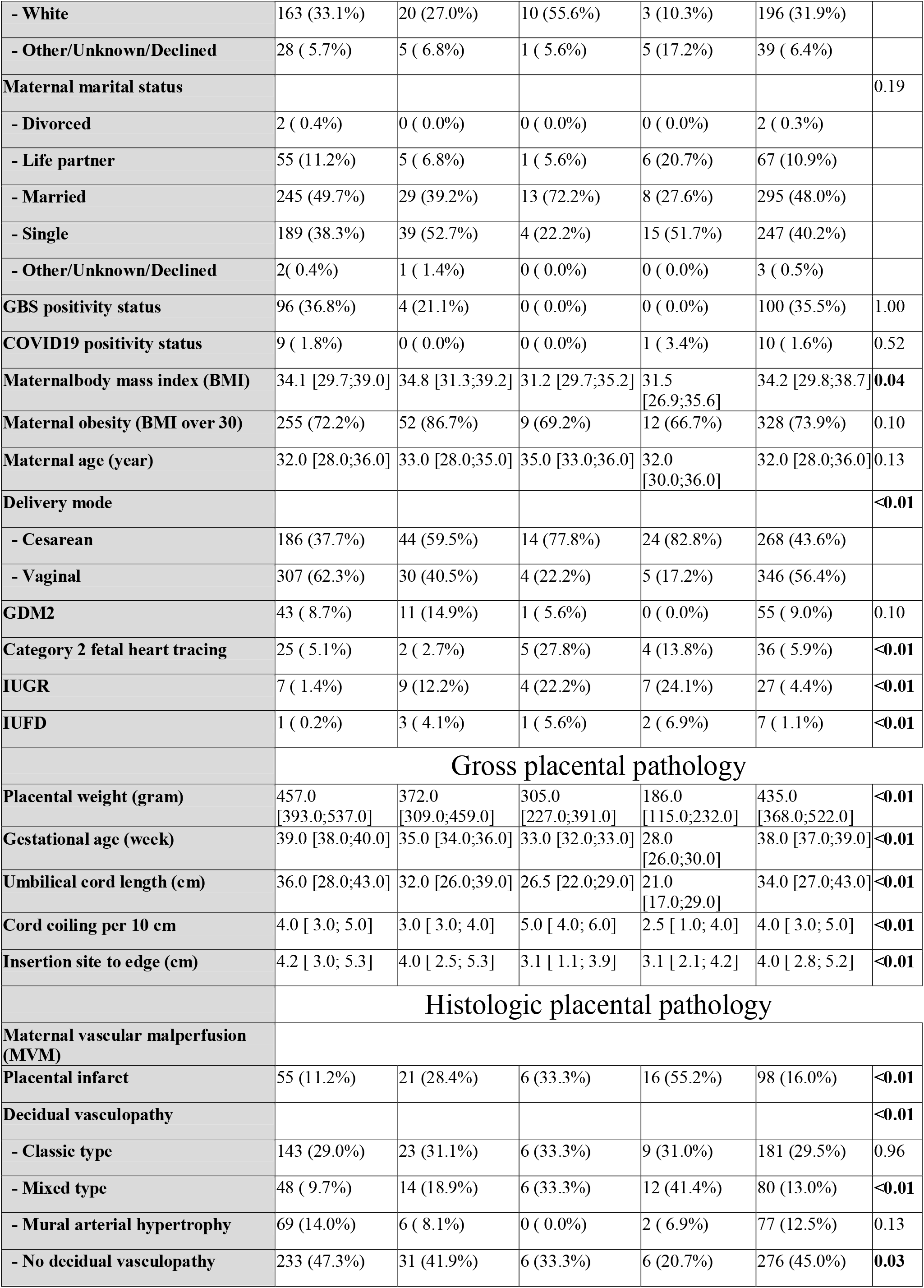
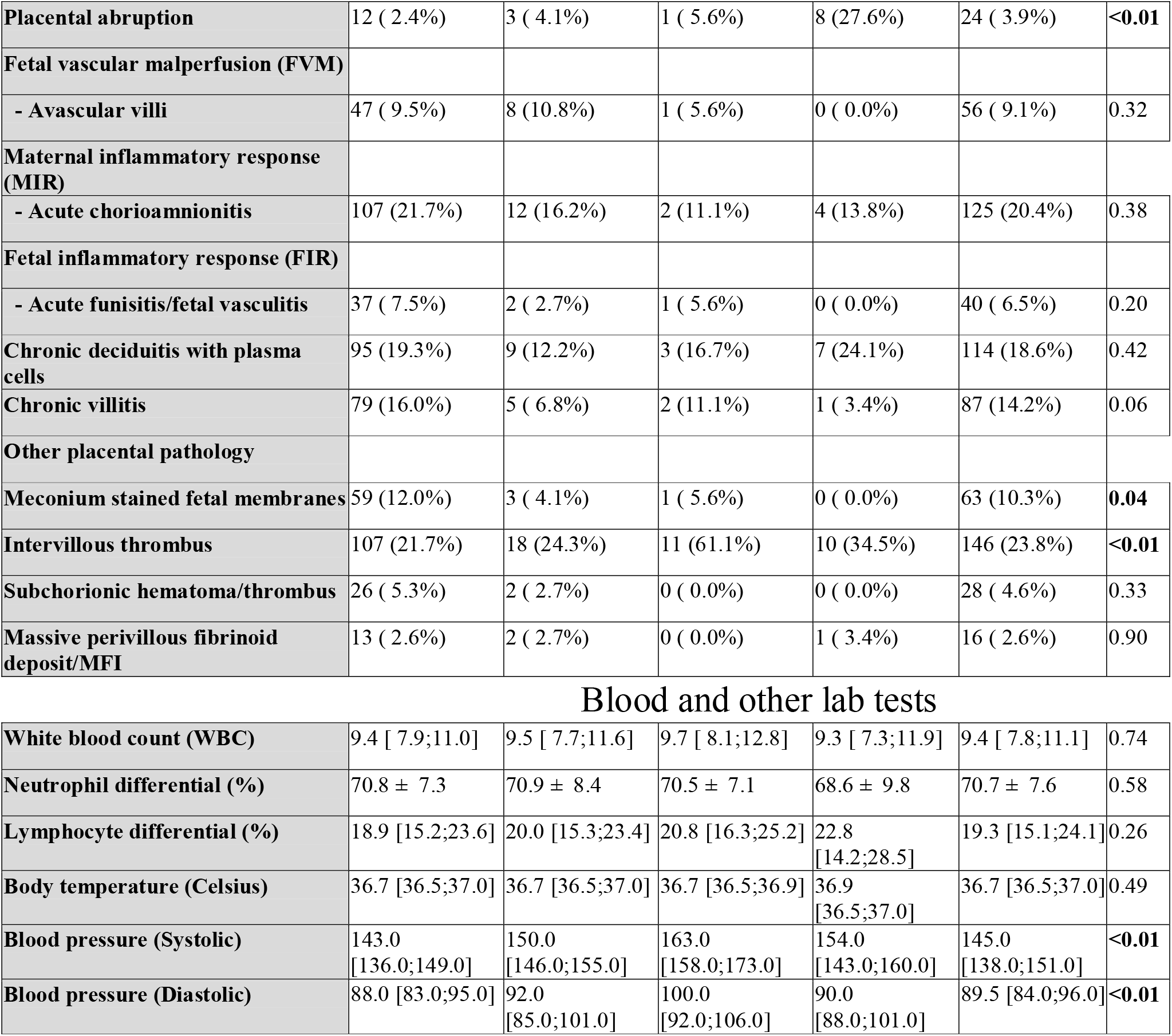
Premature delivery in patients with preeclampsia/PIH.

|  | **Term** | **Late preterm** | **Moderate preterm** | **Extreme preterm** | **Total** | **p** |
| --- | --- | --- | --- | --- | --- | --- |
|  | (N=493)(80.3%) | (N=74)(12.1%) | (N=18)(2.9%) | (N=29)(4.7%) | (N=614) |  |
|  | **Neonatal birth characteristics** |  |  |  |  |  |
| **Neonatal sex** |  |  |  |  |  |  |
| - Female | 258 (52.3%) | 32 (43.2%) | 9 (50.0%) | 15 (51.7%) | 313 (51.0%) |  |
| - Male | 235 (47.7%) | 42 (56.8%) | 9 (50.0%) | 14 (48.3%) | 299 (49.0%) |  |
| **Fetal birth weight (gram)** | 3160.0<br>[2790.0;3490.0] | 2290.0<br>[1835.0;2745.0] | 1670.0<br>[1510.0;1860.0] | 1040.0<br>[560.0;1350.0] | 3010.0<br>[2590.0;3410.0] | **<0.01** |
| **Fetal birth length (cm)** | 50.0 [48.5;51.5] | 46.2 [43.0;48.0] | 42.5 [41.0;43.5] | 36.2<br>[30.0;40.5] | 49.5 [47.0;51.0] | **<0.01** |
| **Fetal head circumference (cm)** | 34.0 [33.0;35.0] | 32.0 [31.0;33.0] | 29.0 [28.5;30.8] | 26.0<br>[22.5;27.5] | 33.5 [32.0;34.5] | **<0.01** |
| **Placental weight (gram)** | 457.0<br>[393.0;537.0] | 372.0<br>[309.0;459.0] | 305.0<br>[227.0;391.0] | 186.0<br>[115.0;232.0] | 435.0<br>[368.0;522.0] | **<0.01** |
| **Fetal placental ratio (FPR)** | 6.9 [ 6.2; 7.6] | 6.0 [ 5.3; 6.7] | 5.4 [ 4.8; 6.5] | 4.7 [ 3.6; 6.3] | 6.7 [ 5.9; 7.6] | **<0.01** |
|  | **Maternal pregnancy characteristics** |  |  |  |  |  |
| **Maternal race /ethnicity** |  |  |  |  |  | 0.09 |
| - Asian | 16 ( 3.2%) | 2 ( 2.7%) | 0 ( 0.0%) | 0 ( 0.0%) | 18 ( 2.9%) |  |
| - Black | 241 (48.9%) | 41 (55.4%) | 6 (33.3%) | 19 (65.5%) | 307 (50.0%) |  |
| - Hispanic | 45 ( 9.1%) | 6 ( 8.1%) | 1 ( 5.6%) | 2 ( 6.9%) | 54 ( 8.8%) |  |
| - White | 163 (33.1%) | 20 (27.0%) | 10 (55.6%) | 3 (10.3%) | 196 (31.9%) |  |
| - Other/Unknown/Declined | 28 ( 5.7%) | 5 ( 6.8%) | 1 ( 5.6%) | 5 (17.2%) | 39 ( 6.4%) |  |
| Maternal marital status |  |  |  |  |  | 0.19 |
| - Divorced | 2 ( 0.4%) | 0 ( 0.0%) | 0 ( 0.0%) | 0 ( 0.0%) | 2 ( 0.3%) |  |
| - Life partner | 55 (11.2%) | 5 ( 6.8%) | 1 ( 5.6%) | 6 (20.7%) | 67 (10.9%) |  |
| - Married | 245 (49.7%) | 29 (39.2%) | 13 (72.2%) | 8 (27.6%) | 295 (48.0%) |  |
| - Single | 189 (38.3%) | 39 (52.7%) | 4 (22.2%) | 15 (51.7%) | 247 (40.2%) |  |
| - Other/Unknown/Declined | 2( 0.4%) | 1 ( 1.4%) | 0 ( 0.0%) | 0 ( 0.0%) | 3 ( 0.5%) |  |
| GBS positivity status | 96 (36.8%) | 4 (21.1%) | 0 ( 0.0%) | 0 ( 0.0%) | 100 (35.5%) | 1.00 |
| COVID19 positivity status | 9 ( 1.8%) | 0 ( 0.0%) | 0 ( 0.0%) | 1 ( 3.4%) | 10 ( 1.6%) | 0.52 |
| Maternalbody mass index (BMI) | 34.1 [29.7;39.0] | 34.8 [31.3;39.2] | 31.2 [29.7;35.2] | 31.5 [26.9;35.6] | 34.2 [29.8;38.7] | **0.04** |
| Maternal obesity (BMI over 30) | 255 (72.2%) | 52 (86.7%) | 9 (69.2%) | 12 (66.7%) | 328 (73.9%) | 0.10 |
| Maternal age (year) | 32.0 [28.0;36.0] | 33.0 [28.0;35.0] | 35.0 [33.0;36.0] | 32.0 [30.0;36.0] | 32.0 [28.0;36.0] | 0.13 |
| Delivery mode |  |  |  |  |  | **<0.01** |
| - Cesarean | 186 (37.7%) | 44 (59.5%) | 14 (77.8%) | 24 (82.8%) | 268 (43.6%) |  |
| - Vaginal | 307 (62.3%) | 30 (40.5%) | 4 (22.2%) | 5 (17.2%) | 346 (56.4%) |  |
| GDM2 | 43 ( 8.7%) | 11 (14.9%) | 1 ( 5.6%) | 0 ( 0.0%) | 55 ( 9.0%) | 0.10 |
| Category 2 fetal heart tracing | 25 ( 5.1%) | 2 ( 2.7%) | 5 (27.8%) | 4 (13.8%) | 36 ( 5.9%) | **<0.01** |
| IUGR | 7 ( 1.4%) | 9 (12.2%) | 4 (22.2%) | 7 (24.1%) | 27 ( 4.4%) | **<0.01** |
| IUFD | 1 ( 0.2%) | 3 ( 4.1%) | 1 ( 5.6%) | 2 ( 6.9%) | 7 ( 1.1%) | **<0.01** |
|  | Gross placental pathology |  |  |  |  |  |
| Placental weight (gram) | 457.0 [393.0;537.0] | 372.0 [309.0;459.0] | 305.0 [227.0;391.0] | 186.0 [115.0;232.0] | 435.0 [368.0;522.0] | **<0.01** |
| Gestational age (week) | 39.0 [38.0;40.0] | 35.0 [34.0;36.0] | 33.0 [32.0;33.0] | 28.0 [26.0;30.0] | 38.0 [37.0;39.0] | **<0.01** |
| Umbilical cord length (cm) | 36.0 [28.0;43.0] | 32.0 [26.0;39.0] | 26.5 [22.0;29.0] | 21.0 [17.0;29.0] | 34.0 [27.0;43.0] | **<0.01** |
| Cord coiling per 10 cm | 4.0 [ 3.0; 5.0] | 3.0 [ 3.0; 4.0] | 5.0 [ 4.0; 6.0] | 2.5 [ 1.0; 4.0] | 4.0 [ 3.0; 5.0] | **<0.01** |
| Insertion site to edge (cm) | 4.2 [ 3.0; 5.3] | 4.0 [ 2.5; 5.3] | 3.1 [ 1.1; 3.9] | 3.1 [ 2.1; 4.2] | 4.0 [ 2.8; 5.2] | **<0.01** |
|  | Histologic placental pathology |  |  |  |  |  |
| Maternal vascular malperfusion (MVM) |  |  |  |  |  |  |
| Placental infarct | 55 (11.2%) | 21 (28.4%) | 6 (33.3%) | 16 (55.2%) | 98 (16.0%) | **<0.01** |
| Decidual vasculopathy |  |  |  |  |  | **<0.01** |
| - Classic type | 143 (29.0%) | 23 (31.1%) | 6 (33.3%) | 9 (31.0%) | 181 (29.5%) | 0.96 |
| - Mixed type | 48 ( 9.7%) | 14 (18.9%) | 6 (33.3%) | 12 (41.4%) | 80 (13.0%) | **<0.01** |
| - Mural arterial hypertrophy | 69 (14.0%) | 6 ( 8.1%) | 0 ( 0.0%) | 2 ( 6.9%) | 77 (12.5%) | 0.13 |
| - No decidual vasculopathy | 233 (47.3%) | 31 (41.9%) | 6 (33.3%) | 6 (20.7%) | 276 (45.0%) | **0.03** |
| **Placental abruption** | 12 ( 2.4%) | 3 ( 4.1%) | 1 ( 5.6%) | 8 (27.6%) | 24 ( 3.9%) | **<0.01** |
| **Fetal vascular malperfusion (FVM)** |  |  |  |  |  |  |
| - Avascular villi | 47 ( 9.5%) | 8 (10.8%) | 1 ( 5.6%) | 0 ( 0.0%) | 56 ( 9.1%) | 0.32 |
| **Maternal inflammatory response (MIR)** |  |  |  |  |  |  |
| - Acute chorioamnionitis | 107 (21.7%) | 12 (16.2%) | 2 (11.1%) | 4 (13.8%) | 125 (20.4%) | 0.38 |
| **Fetal inflammatory response (FIR)** |  |  |  |  |  |  |
| - Acute funisitis/fetal vasculitis | 37 ( 7.5%) | 2 ( 2.7%) | 1 ( 5.6%) | 0 ( 0.0%) | 40 ( 6.5%) | 0.20 |
| **Chronic deciduitis with plasma cells** | 95 (19.3%) | 9 (12.2%) | 3 (16.7%) | 7 (24.1%) | 114 (18.6%) | 0.42 |
| **Chronic villitis** | 79 (16.0%) | 5 ( 6.8%) | 2 (11.1%) | 1 ( 3.4%) | 87 (14.2%) | 0.06 |
| **Other placental pathology** |  |  |  |  |  |  |
| **Meconium stained fetal membranes** | 59 (12.0%) | 3 ( 4.1%) | 1 ( 5.6%) | 0 ( 0.0%) | 63 (10.3%) | **0.04** |
| **Intervillous thrombus** | 107 (21.7%) | 18 (24.3%) | 11 (61.1%) | 10 (34.5%) | 146 (23.8%) | **<0.01** |
| **Subchorionic hematoma/thrombus** | 26 ( 5.3%) | 2 ( 2.7%) | 0 ( 0.0%) | 0 ( 0.0%) | 28 ( 4.6%) | 0.33 |
| **Massive perivillous fibrinoid deposit/MFI** | 13 ( 2.6%) | 2 ( 2.7%) | 0 ( 0.0%) | 1 ( 3.4%) | 16 ( 2.6%) | 0.90 |
| Blood and other lab tests |  |  |  |  |  |  |
| **White blood count (WBC)** | 9.4 [ 7.9;11.0] | 9.5 [ 7.7;11.6] | 9.7 [ 8.1;12.8] | 9.3 [ 7.3;11.9] | 9.4 [ 7.8;11.1] | 0.74 |
| **Neutrophil differential (%)** | 70.8 ± 7.3 | 70.9 ± 8.4 | 70.5 ± 7.1 | 68.6 ± 9.8 | 70.7 ± 7.6 | 0.58 |
| **Lymphocyte differential (%)** | 18.9 [15.2;23.6] | 20.0 [15.3;23.4] | 20.8 [16.3;25.2] | 22.8 [14.2;28.5] | 19.3 [15.1;24.1] | 0.26 |
| **Body temperature (Celsius)** | 36.7 [36.5;37.0] | 36.7 [36.5;37.0] | 36.7 [36.5;36.9] | 36.9 [36.5;37.0] | 36.7 [36.5;37.0] | 0.49 |
| **Blood pressure (Systolic)** | 143.0 [136.0;149.0] | 150.0 [146.0;155.0] | 163.0 [158.0;173.0] | 154.0 [143.0;160.0] | 145.0 [138.0;151.0] | **<0.01** |
| **Blood pressure (Diastolic)** | 88.0 [83.0;95.0] | 92.0 [85.0;101.0] | 100.0 [92.0;106.0] | 90.0 [88.0;101.0] | 89.5 [84.0;96.0] | **<0.01** |

### 2, Preterm delivery and clinical pathologic features in patients with PRE/PIH

There were 614 placentas from patients with preeclampsia/PIH (PRE/PIH), and these cases were divided as term (n=493), late preterm (n=74), moderate preterm (n=18) and extreme preterm (n=29). PRE/PIH is known to be associated with iatrogenic preterm deliveries and significant placental pathology features as described previously (Table 2). As expected, PRE/PIH was significantly associated with progressive lower neonatal birth weight, birth length, head circumference, placental weight and FPR (p<0.01 for all, Table 2). PRE/PIH was significantly associated with increased C-section delivery, category 2 fetal heart tracing, IUGR and IUFD (p<0.01 for all). PRE/PIH was significantly associated with features of maternal vascular malperfusion (MVM), characterized by the presence of placental infarct, decidual vasculopathy, placental abruption and intervillous thrombosis (p<0.01 for all, Table 2). PRE/PIH was significantly associated with elevated systolic and diastolic blood pressure at the time of delivery (p<0.01, Table 2).

### 3, Preterm delivery and clinical pathologic features in patients without PRE/PIH

There were 3,109 placental cases from patients without PRE/PIH, categorized as term (≥37 weeks, n=2,814), late preterm (34–36 weeks, n=184), moderate preterm (32–33 weeks, n=20), and extreme preterm (<32 weeks, n=91) (Table 3). Preterm delivery groups showed progressively lower neonatal birth weight (median 3,310 g term vs. 810 g extreme preterm), placental weight (466 g vs. 194 g), and FPR (7.1 vs. 4.2) (p<0.01 for all, Table 3). Ethnic black maternal race was enriched in preterm groups (30.4% term vs. 46.2% extreme preterm, p<0.01). Intra-amniotic infection including maternal inflammatory response (acute chorioamnionitis) (36.1% term vs. 70.3% extreme preterm) and fetal inflammatory response (funisitis and fetal vasculitis, 14.3% vs. 38.5%) were significantly higher in preterm deliveries (p<0.01, Table 3). Intrauterine fetal demise was markedly elevated in extreme preterm (34.1% vs. 0.2% term, p<0.01). In addition, WBC was elevated in preterm delivery groups and so was the body temperature at the time of delivery (p<0.01 and p=0.01, Table 3).

**Table 3:**
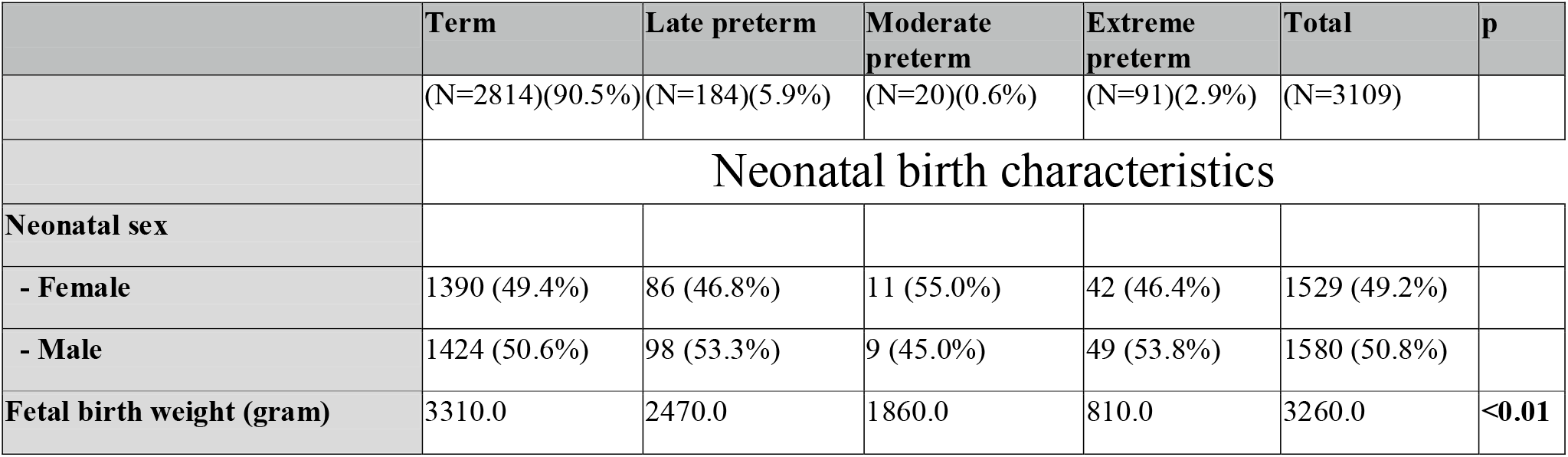
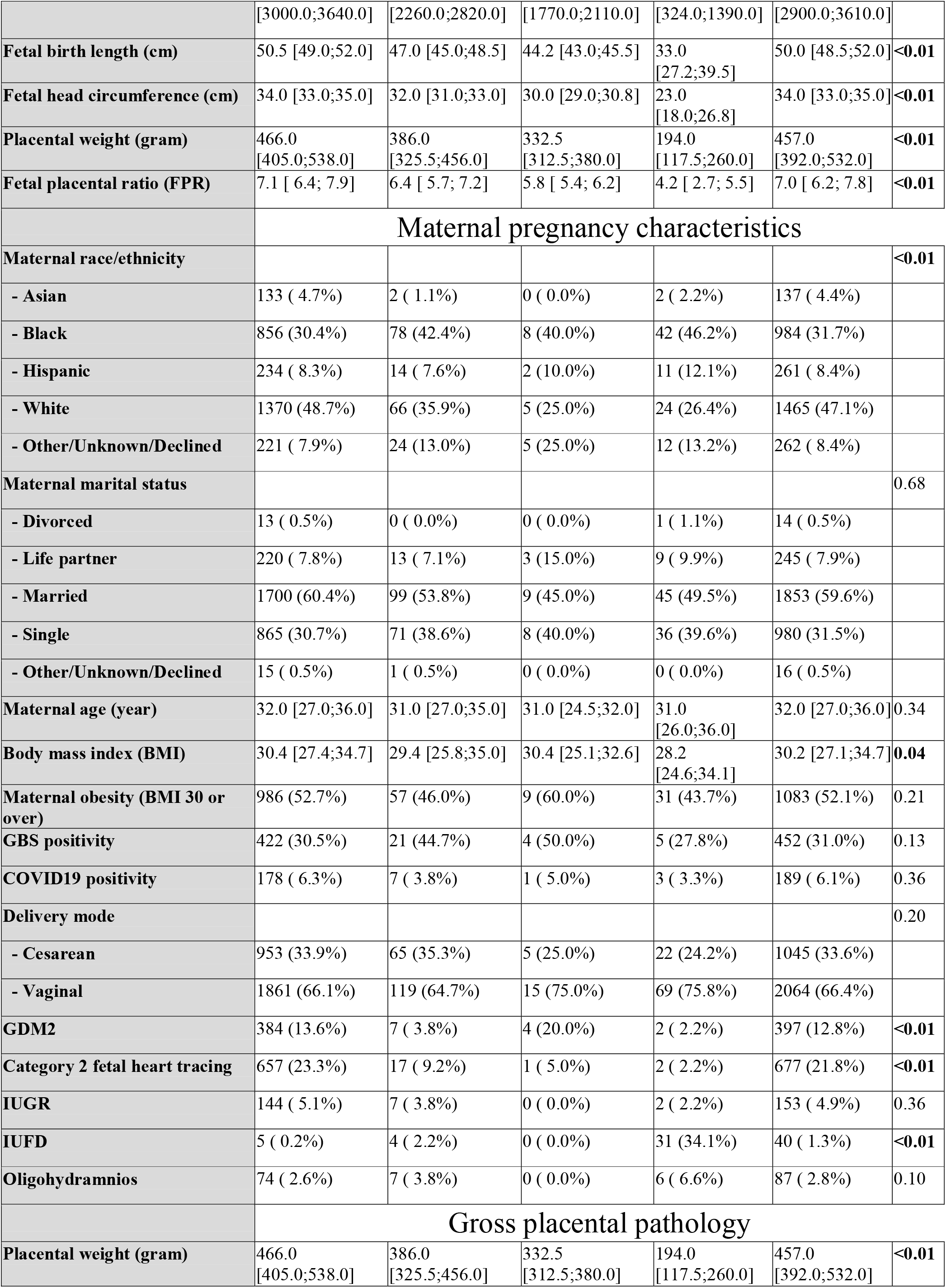
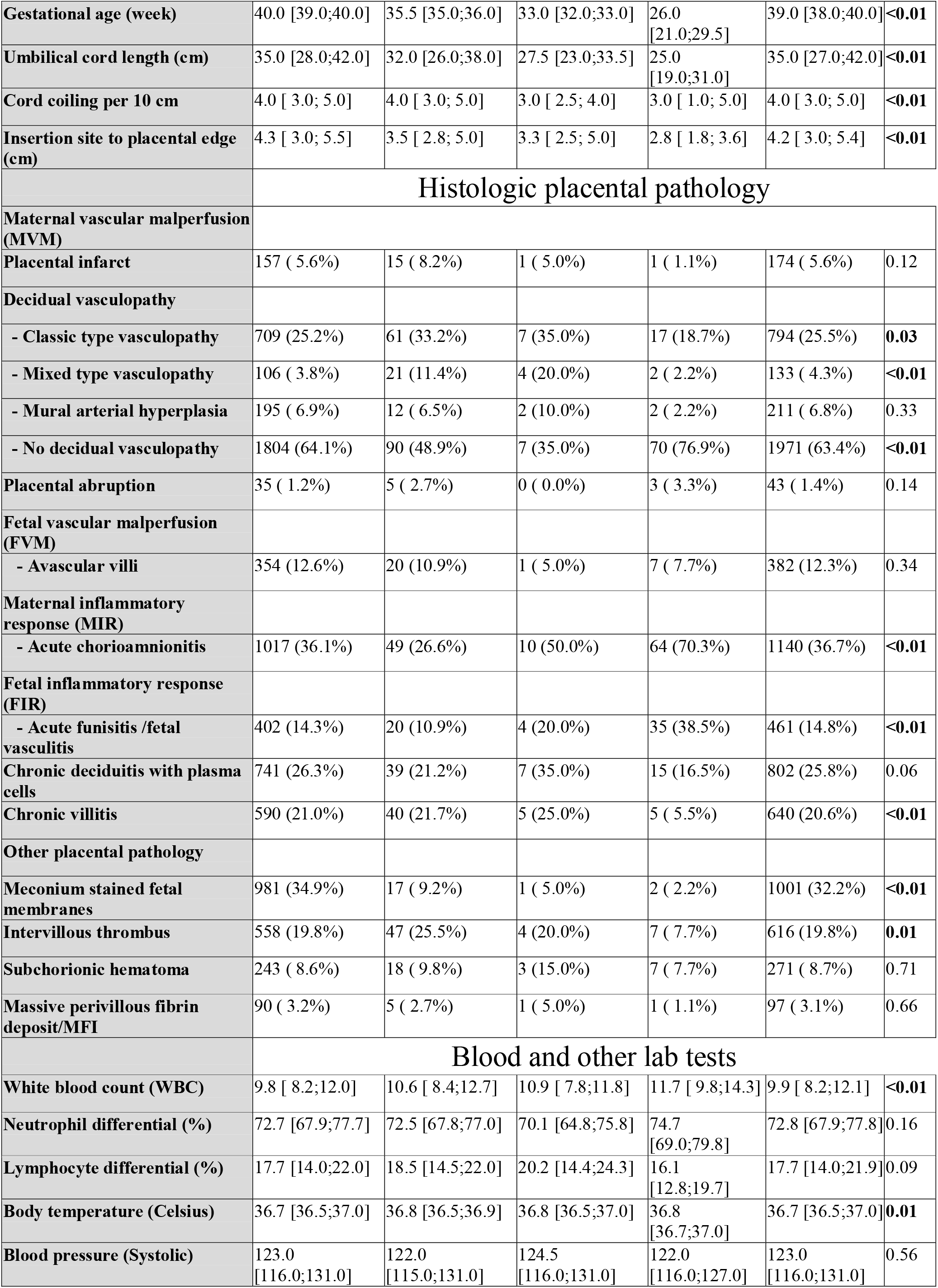
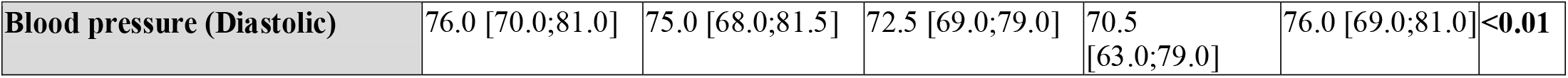
Preterm delivery and clinicopathologic features without PRE/PIH.

## Discussion

Preterm delivery has been previously classified as spontaneous type and iatrogenic type with distinct pathogenic mechanisms and clinical management ^9^. Spontaneous preterm delivery was more commonly associated with intra-amniotic infection while iatrogenic type was more commonly associated with hypertensive disorders of pregnancy/preeclampsia ^9^. Our cohort data demonstrated clear clinicopathologic distinctions across preterm subcategories. Maternal vascular-related pathologic features (preeclampsia, MVM lesions, higher blood pressure) were enriched in late and moderate preterm deliveries in patients with PRE/PIH, consistent with established associations between hypertensive disorders of pregnancy and related preterm birth ^10,11^. The progressive decrease in fetal and placental weight reflects expected growth restrictions in earlier gestations due to maternal vascular malperfusion. In patient populations without PRE/PIH, the markedly elevated acute chorioamnionitis and fetal inflammatory response strongly support infection/inflammation as a dominant mechanism in non-hypertensive preterm delivery^10,12^. This is consistent with historic literature linking chorioamnionitis or intra-amniotic infection to spontaneous preterm delivery ^13-15^. Extreme preterm deliveries (<32 weeks) were associated with strikingly high rates of intrauterine fetal death (IUFD, 27.5%) and acute histologic chorioamnionitis/funisitis (maternal and fetal inflammatory response)(56.7%/29.2%), suggesting that intra-amniotic infection is a significant factor in this subgroup with implication for clinical management ^16,17^.

Racial disparities were evident, with ethnic black mothers significantly overrepresented in preterm groups - a finding consistent with extensive epidemiologic studies on social, biologic, and healthcare factors contributing to higher preterm birth rates in black population ^5,15,18-21^.

The strengths of our study include the large sample size, detailed systematic placental pathology examination, and comprehensive clinical data. Limitations include the retrospective design, potential single-center bias, lack of multivariate adjustment for confounders, and absence of long-term neonatal outcomes.

These results reinforce the heterogeneous nature of preterm deliveries and the diagnostic value of placental examination in elucidating underlying mechanisms.

## Conclusion

Preterm delivery is associated with distinct clinicopathologic profiles. Iatrogenic preterm delivery was significantly associated with maternal vascular malperfusion in late preterm categories, while spontaneous extreme preterm cases frequently involved intrauterine fetal demise and intra-amniotic infection (maternal and fetal inflammatory response). Placental examination provides important information which may improve etiologic understanding and guide future preventive strategies.

## Data Availability

All data produced in the present work are contained in the manuscript.

## Financial disclosure

The authors declare no financial conflict of interest.

## Funding disclosure

The study received no funding.

## Ethnic statement

This study and placental data collection was approved by the Institutional Review Board of New York Presbyterian –Brooklyn Methodist Hospital [1592673-1] (approval date 4-13-2020).

## Notes

### Competing Interest Statement

The authors have declared no competing interest.

### Funding Statement

This study did not receive any funding.

### Author Declarations

The placental data collection was approved by the Institutional Review Board of New York Presbyterian Brooklyn Methodist Hospital [15926731] (approval date 4/13/2020).

